# Research Involvement and Partnership Outcomes Among Patient-Centered Outcomes Research Partners

**DOI:** 10.64898/2026.11.14.26368107

**Authors:** Vedant N. Tapiavala, Jemar R. Bather, Emma Ricci-De Lucca, Elizabeth L. Murnane, Melody S. Goodman, Karen L. Fortuna

## Abstract

**Importance:** Meaningful patient and community involvement is increasingly emphasized in health research. Still, little is known about how such research involvement relates to the quality of patient and community partnerships.

**Objective:** To examine the associations of patient and community involvement in research with partnership outcomes. The associations of patient involvement in specific research phases (e.g., grant writing, recruitment, and data collection) with these metrics were examined as a secondary objective.

**Design:** This cross-sectional observational study used baseline data from a national cohort study across the United States collected from September to December 2024.

**Setting:** This national study was conducted online with recruitment occurring over multiple channels.

**Participants:** Participants included patient and community research partners aged 18 years and older and were recruited in a convenience sample.

**Exposures:** The extent to which participants have contributed to prior research studies were calculated by adding binary indicators representing their involvement in phases of the research process. These indicators were examined individually as secondary exposures.

**Main Outcome(s) and Measure(s):** Primary outcomes included partnership quality and research engagement, with trust in medical researchers and partnership recommendation as secondary and tertiary outcomes. Multivariable linear and modified Poisson regressions were utilized to estimate regression coefficients and adjusted prevalence ratios (aPRs).

**Results:** Among 330 participants (mean age = 36.5 years, SD = 9.7), 56.1% were male, and 37.3% and 31.8% were White or Black, respectively. Involvement in patient-centered outcomes research was associated with partnership quality (β, 0.86; 95% CI, 0.30-1.42, P=0.003), research engagement (β, 0.04; 95% CI, 0.01-0.06, P=0.006), trust in medical researchers (β, 0.69; 95% CI, 0.31-1.07, P<0.001), and greater willingness to recommend becoming a research partner (aPR, 1.09; 95% CI, 1.05-1.14, P<0.001).

**Conclusions and Relevance:** Patient-centered outcomes research may provide patient and community research partners with a sense of shared ownership, improving partnership quality. While the causal links require further investigation, these associations may be driven by inclusion in recruitment, grant writing, data collection, and shared governance.

**Key Points:**

1. Is greater patient involvement in patient-centered outcomes research associated with partnership quality, research engagement, trust in medical researchers, and partnership recommendation?
2. This national cohort study found that patient research involvement was significantly associated with all four primary outcomes and that these associations were primarily driven by patient partners’ participation in recruitment, grant writing, data collection, and shared governance.
3. Patient-centered outcomes research involvement across the research process may improve partnership quality, indicating the importance of patient and community partnership in shared decision-making and roles creating visible impact.

## Introduction

Patient-centered outcomes research offers patients and community members a pathway to offer their lived experience expertise to shape research projects.^1^ Patient and community involvement in such research can span the entire research process, from ideation and grant writing to defining research questions, identifying meaningful outcomes, interpreting findings, and disseminating results.^2,3^ Patient-centered outcomes research may increase representation of underserved populations in research,^4,5^ improve research engagement and partnership quality,^6^ and align decision-making with patients’ perspectives and experiences.^7^ Engaging patients and communities as partners in research has been linked to more relevant interventions and better uptake of findings in the real world beyond clinical settings,^8,9^ which could shorten the gap between developing an intervention and adopting it into practice.^10,11^

Evidence for these benefits, however, remains limited. A recent systematic review of 49 studies found that engagement strategies ranged from consultation to active involvement in planning, execution, and evaluation.^11^ Definitions and outcome measures varied widely with no shared reporting structure, so it remains unclear whether engagement accelerates the translation of research into practice. Trust is considered central to patient engagement,^12^ but empirical findings on whether engagement builds trust in research are limited and mixed. One 12-month community stakeholder group consisting of 12 Black community leaders reported improved trust in engagement with research,^13^ whereas 13 participants in a single-arm pre-post pilot study reported greater distrust of the medical system after training in community-based participatory research.^6^

Patient-centered outcomes research has become a funding priority in health research, as evidenced by the Patient-Centered Outcomes Research Institute (PCORI) requiring meaningful patient and stakeholder engagement in the research it funds^14,15^ and the NIH Community Engagement Alliance partnering with nearly 1,000 organizations nationwide.^16^ Despite this growing recognition of the importance of meaningful patient and community engagement, little is known about whether greater patient and community involvement in research partnerships is associated with higher partnership quality, greater research engagement, increased trust in medical researchers, and a greater willingness to recommend serving as a research partner. Understanding these associations is needed for identifying appropriate engagement strategies that support high-quality partnerships, build trust, and sustain meaningful stakeholder engagement throughout the research process.

This study examined the associations of research involvement with the outcomes of patient and community partnerships. Using data from a national cohort of patient and community research partners (N = 330), we examined whether greater research involvement was associated with (1) partnership quality, (2) research engagement, (3) trust in medical researchers, and (4) willingness to recommend being a research partner. We additionally explored whether specific types of research involvement, such as study design, recruitment, and shared governance, were differentially associated with these outcomes.

## Methods

### Study Design, Setting, and Sample

This cross-sectional study used baseline data from a cohort study of patient and community research partners in the United States. Data were primarily collected to validate the Quality of Patient-Centered Outcomes Research Partnerships (QPCOR) instrument,^17^ and this analysis represents a secondary use of that data. Eligibility criteria required participants to be at least 18 years old and involved in patient-centered outcomes research as a patient or community partner. Eligible participants were asked to complete an online survey administered at study enrollment and at three- and six-month follow-up engagements, though this analysis is restricted to baseline data collected at study enrollment. Secondary measures included research involvement and engagement, trust in medical researchers, willingness to recommend becoming a research partner, and sociodemographic factors.

A total of 330 participants enrolled between September and December 2024. Participants were recruited nationally through multiple channels, including emails to active PCORI investigators, social media (e.g., LinkedIn posts), universally designed recruitment flyers,^18^ newsletters, listserv announcements, and personal referrals. Given the online recruitment approach, a fraud prevention protocol was implemented in which research staff conducted a 15- minute virtual session with each prospective participant to verify identity, project affiliation, and collaborating investigator. All participants provided informed consent, and all study procedures were approved by the Dartmouth College Institutional Review Board. Reporting follows the Strengthening the Reporting of Observational Studies in Epidemiology guidelines.^19^

### Outcome Measures

#### Quality of Patient-Centered Outcomes Research Partnerships

The first primary outcome was quality of patient-centered outcomes research partnerships (hereafter, partnership quality) assessed using the 10-item QPCOR instrument.^6,17^ Participants were presented with the following prompt: “Consider your Patient-Centered Outcomes Research partnerships to date in one specific project and respond to each of the following questions on a scale from *0 = No effort was made by researchers* to *10 = Every effort was made by researchers*.” The scale consisted of ten items, which can be found in Appendix A. A composite score was constructed as the sum of the ten items, such that scores could range from 0 to 100, with higher scores reflecting greater partnership quality (Cronbach’s α = 0.94).^20^ The QPCOR development process is described in detail elsewhere.^6,17^

#### Research Engagement

The second primary outcome was research engagement measured using the Brief Research Engagement Survey Tool^21^ to complement the quality-focused metrics of the QPCOR instrument. Using a 5-point Likert scale ranging from 1 *(never)* to 5 *(always)*, respondents rated how often the partners leading the research engaged in one of nine behaviors, which are listed in Appendix A. A composite score was derived as the mean of the nine responses, with higher values reflecting greater research engagement (Cronbach’s α = 0.86).

#### Trust in Medical Researchers

The secondary outcome of trust in medical researchers was assessed using the 12-item Trust in Medical Researchers Scale.^22^ Using a 5-point Likert scale ranging from 1 *(strongly disagree)* to 5 *(strongly agree)*, respondents rated each item, all of which are detailed in Appendix A. Prior to constructing subscale scores, items 1-6, 11, and 12 were reverse-coded. A total scale score was derived as the sum of all 12 items, minus the minimum possible score of 12, such that scores ranged from 0 to 48, with higher scores reflecting greater trust in medical researchers (Cronbach’s α = 0.86).

#### Research Partner Recommendation

The tertiary outcome of partnership recommendation was assessed using one item. Participants were asked: “On a scale from 1-10, how likely are you to recommend being a research partner? (1 = not at all, 10 = highly likely)”. To distinguish those who strongly recommended being a research partner from those who did not, responses were dichotomized as *highly likely (1)* versus *all other responses (0)*.

### Exposure Ascertainment

#### Research Involvement

Research involvement was measured using the following item: “In what ways have you contributed to your research study?” Participants could select all that applied from the following responses: (1) study design, (2) recruitment, (3) retention management, (4) data interpretation, (5) dissemination, (6) grant writing, (7) community advisory board, (8) data collection, (9) shared governance, and (10) other. A composite measure (primary exposure) was constructed as the sum of checked responses. As secondary exposures, we created separate indicator variables for each item (1 = Yes, 0 = No).

### Covariates

Covariates included age (<30 years, 30 to 39 years, 40 to 49 years, 50 years or older), sex (male, female, non-Binary/transgender), race/ethnicity (White, Black, Hispanic, Other), marital status (married, single, divorced/separated), educational attainment (high school/occupational training, some college/associate’s degree, college degree or higher), and census region (Northeast, South, Midwest, West). The Other racial/ethnic category comprised participants identifying as Asian, American Indian/Alaskan Native, Native Hawaiian/Pacific Islander, or Multiracial. The U.S. census region was derived from self-reported state of residence.

### Statistical Analysis

Descriptive statistics were expressed as counts and proportions for categorical variables and as means and SDs for continuous measures. Multivariable linear regression was used to estimate the adjusted associations of research involvement with partnership quality, research engagement, and trust in medical researchers.^23^ For partnership recommendation, we employed modified Poisson regression with generalized estimating equations to estimate adjusted prevalence ratios (aPRs) with 95% confidence intervals (CIs).^24,25^ This approach avoids convergence issues associated with log-binomial regression and yields the PR as a more interpretable association measure than the odds ratio.^26,27^ An independence working correlation structure was specified. All models adjusted for age, sex, race/ethnicity, marital status, educational attainment, and census region. Given the descriptive and exploratory nature of this study, adjustment for multiple comparisons was not applied to reduce the risk of Type II error.^28^ A two-sided *p*<0.05 was considered statistically significant.

A total of 52 participants (15.8%) had incomplete data across seven distinct missing data patterns. Little’s Missing Completely at Random (MCAR) test indicated that missing data were not MCAR ( = 135.20, *p*<0.001).^29^ As such, multiple imputation by chained equations (mice package in R) was used to address missing data under a Missing at Random assumption.^30^ Imputation models were specified according to variable type: predictive mean matching for research engagement, ordinal logistic regression for educational attainment, and multinomial logistic regression for census region. Results were pooled across 15 imputed datasets using Rubin’s rules.^31^ All statistical analyses were performed in R version 4.5.2 (R Core Team, R Foundation for Statistical Computing).

## Results

### Descriptive statistics

Characteristics of the 330 participants are presented in Table 1. Nearly three-quarters (72.4%) were at least 30 years old, 56.1% were male, 56.4% were married, 48.2% had a college degree or higher, and 39.4% resided in the South census region. The racial/ethnic distribution was 37.3% White, 31.8% Black, 10.9% Hispanic, and 20.0% Other. Regarding research involvement, participants contributed to approximately three research activities on average (mean = 2.6, SD = 2.5). The highest involvement was in recruitment (43.6%), followed by data collection (36.7%), dissemination (32.7%), and data interpretation (29.1%). On average, participants reported high partnership quality (mean = 86.7, SD = 12.6), high research engagement (mean = 4.3, SD = 0.5), and moderate trust in medical researchers (mean = 28.2, SD = 8.1). Forty-five percent indicated that they would highly recommend being a research partner.

**Table 1.** Descriptive statistics from a national sample of patient and community research partners in the United States.

| **Characteristic** | **N = 330** |
|---|---|
| Age, n (%) |  |
| <30 | 91 (27.6) |
| 30-39 | 137 (41.5) |
| 40-49 | 71 (21.5) |
| 50+ | 31 (9.4) |
| Sex, n (%) |  |
| Male | 185 (56.1) |
| Female | 125 (37.9) |
| non-Binary/Transgender | 20 (6.1) |
| Race/ethnicity, n (%) |  |
| White | 123 (37.3) |
| Black | 105 (31.8) |
| Hispanic | 36 (10.9) |
| Other | 66 (20.0) |
| Marital status, n (%) |  |
| Married | 186 (56.4) |
| Single | 102 (30.9) |
| Divorced/Separated | 42 (12.7) |
| Educational attainment, n (%) |  |
| High school/Occupational training | 23 (7.0) |
| Some college/Associate’s degree | 109 (33.0) |
| College degree or higher | 159 (48.2) |
| Missing | 39 (11.8) |
| Census region, n (%) |  |
| Northeast | 73 (22.1) |
| South | 130 (39.4) |
| Midwest | 30 (9.1) |
| West | 83 (25.2) |
| Missing | 14 (4.2) |
| Research involvement, Mean (SD) | 2.6 (2.5) |
| Study design, n (%) | 82 (24.8) |
| Recruitment, n (%) | 144 (43.6) |
| Retention management, n (%) | 70 (21.2) |
| Data interpretation, n (%) | 96 (29.1) |
| Dissemination, n (%) | 108 (32.7) |
| Grant writing, n (%) | 64 (19.4) |
| Community advisory board, n (%) | 94 (28.5) |
| Data collection, n (%) | 121 (36.7) |
| Shared governance, n (%) | 55 (16.7) |
| Other, n (%) | 11 (3.3) |
| Quality of patient-centered outcomes research partnership, Mean (SD) | 86.7 (12.6) |
| Research engagement, Mean (SD) | 4.3 (0.5) |
| Missing, n (%) | 5 (1.5) |
| Trust in medical researchers, Mean (SD) | 28.2 (8.1) |
| Highly recommend being a research partner, n (%) | 148 (44.8) |

### Associations with Research Involvement

The adjusted associations between the composite research involvement score and each outcome are shown in Table 2. Regression models indicated that greater research involvement was significantly associated with higher partnership quality (β = 0.86, 95% CI: 0.30 to 1.42), research engagement (β = 0.04, 95% CI: 0.01 to 0.06), and trust in medical researchers (β = 0.69, 95% CI: 0.31 to 1.07). There was also evidence that greater research involvement was significantly associated with a 9% (aPR = 1.09, 95% CI: 1.05 to 1.14) higher prevalence of highly recommending being a research partner. All associations were adjusted for age, sex, race/ethnicity, marital status, educational attainment, and census region. Full model outputs are displayed in Supplemental Table 1. The associations with partnership quality (p=0.51), research engagement (p=0.49), trust in medical researchers (p=0.30), and partnership recommendation (p=0.67) did not differ by race.

**Table 2.** Adjusted associations of research involvement with partnership quality, research engagement, trust in medical researchers, and partnership recommendation among a U.S. sample of patient-centered outcomes research partners.

|  | **Quality of patient-centered outcomes research partnership** |  | **Research engagement** |  | **Trust in medical researchers** |  | **Highly recommend being a research partner** |  |
|---|---|---|---|---|---|---|---|---|
| **Characteristic** | **b (95% CI)** | **p-value** | **b (95% CI)** | **p-value** | **b (95% CI)** | **p-value** | **aPR (95% CI)** | **p-value** |
| Research involvement | **0.86 (0.30 to 1.42)** | **0.003** | **0.04 (0.01 to 0.06)** | **0.006** | **0.69 (0.31 to 1.07)** | **<0.001** | **1.09 (1.05 to 1.14)** | **<0.001** |

Abbreviations: CI = Confidence Interval, aPR = adjusted Prevalence Ratio
Results were pooled across 15 imputed datasets.
Bold font indicates *p*<0.05.
All models adjusted for age, sex, race/ethnicity, marital status, educational attainment, and census region.

### Associations with Single Research Involvement Indicators

Table 3 presents the adjusted associations between individual research involvement indicators and each outcome. Contributions to recruitment, grant writing, data collection, and shared governance were each significantly associated with higher partnership quality, greater research engagement, increased trust in medical researchers, and stronger partnership recommendation (all *p*<0.05). Involvement in dissemination was significantly associated with partnership quality (β = 4.01, 95% CI: 1.25 to 6.77), research engagement (β = 0.16, 95% CI: 0.04 to 0.28), and partnership recommendation (aPR = 1.36, 95% CI: 1.07 to 1.72), but not with trust in medical researchers (β = 0.44, 95% CI: -1.45 to 2.34). Other statistically significant relationships included data interpretation with trust in medical researchers (β = 2.05, 95% CI: 0.08 to 4.02) and partnership recommendation (aPR = 1.33, 95% CI: 1.02 to 1.71), and retention management with trust in medical researchers (β = 2.36, 95% CI: 0.13 to 4.58). In contrast, no statistically significant associations were observed for study design (all *p*≥0.32) or community advisory board (all *p*≥0.10) contributions with any outcome.

**Table 3.** Adjusted associations of each research involvement indicator with partnership quality, research engagement, trust in medical researchers, and partnership recommendation among a U.S. sample of patient-centered outcomes research partners.

|  | **Quality of patient-centered outcomes research partnership** |  | **Research engagement** |  | **Trust in medical researchers** |  | **Highly recommend being a research partner** |  |
|---|---|---|---|---|---|---|---|---|
| **Exposure model** | **b (95% CI)** | **p-value** | **b (95% CI)** | **p-value** | **b (95% CI)** | **p-value** | **aPR (95% CI)** | **p-value** |
| Study design | -0.71 (-3.80 to 2.39) | 0.65 | -0.01 (-0.15 to 0.12) | 0.87 | 0.87 (-1.23 to 2.97) | 0.42 | 1.15 (0.87 to 1.53) | 0.32 |
| Recruitment | **2.65 (0.09 to 5.21)** | **0.043** | **0.12 (0.01 to 0.24)** | **0.031** | **2.74 (1.02 to 4.45)** | **0.002** | **1.33 (1.06 to 1.68)** | **0.016** |
| Retention management | 1.03 (-2.27 to 4.33) | 0.54 | 0.12 (-0.02 to 0.27) | 0.10 | **2.36 (0.13 to 4.58)** | **0.038** | 1.32 (0.99 to 1.76) | 0.059 |
| Data interpretation | 0.66 (-2.27 to 3.59) | 0.66 | -0.02 (-0.14 to 0.11) | 0.80 | **2.05 (0.08 to 4.02)** | **0.041** | **1.33 (1.02 to 1.71)** | **0.032** |
| Dissemination | **4.01 (1.25 to 6.77)** | **0.005** | **0.16 (0.04 to 0.28)** | **0.011** | 0.44 (-1.45 to 2.34) | 0.64 | **1.36 (1.07 to 1.72)** | **0.012** |
| Grant writing | **4.58 (1.27 to 7.88)** | **0.007** | **0.19 (0.05 to 0.34)** | **0.009** | **3.48 (1.25 to 5.71)** | **0.002** | **1.48 (1.14 to 1.93)** | **0.004** |
| Community advisory board | 0.23 (-2.77 to 3.22) | 0.88 | -0.05 (-0.18 to 0.08) | 0.44 | 1.70 (-0.32 to 3.73) | 0.10 | 1.20 (0.94 to 1.55) | 0.15 |
| Data collection | **4.41 (1.76 to 7.05)** | **0.001** | **0.18 (0.07 to 0.30)** | **0.002** | **2.40 (0.59 to 4.20)** | **0.009** | **1.32 (1.04 to 1.68)** | **0.023** |
| Shared governance | **9.27 (5.73 to 12.81)** | **<0.001** | **0.35 (0.19 to 0.50)** | **<0.001** | **4.66 (2.22 to 7.11)** | **<0.001** | **2.13 (1.65 to 2.75)** | **<0.001** |
| Other | 1.44 (-5.67 to 8.56) | 0.69 | 0.08 (-0.23 to 0.40) | 0.61 | -2.46 (-7.27 to 2.35) | 0.32 | 0.64 (0.21 to 1.97) | 0.43 |

Abbreviations: CI = Confidence Interval, aPR = adjusted Prevalence Ratio
Results were pooled across 15 imputed datasets.
Bold font indicates *p*<0.05.
All models adjusted for age, sex, race/ethnicity, marital status, educational attainment, and census region.

## Discussion

Patient-centered outcomes research may broaden clinical trial participation, strengthen partnerships between researchers and communities, and speed the translation of findings into practice. Still, little is known about the extent to which greater patient involvement in research partnerships is associated with higher partnership quality, greater research engagement, increased trust in medical researchers, and a greater willingness to recommend serving as a research partner. Greater research involvement was significantly associated with higher partnership quality, greater research engagement, increased trust in medical researchers, and stronger partnership recommendation in a national sample of U.S. patient and community research partners. Secondary analyses suggested that these associations may be driven by contributions to recruitment, grant writing, data collection, and shared governance. By contrast, contributions to study design and community advisory boards showed no statistically significant associations with any of the four outcomes. Together, these findings point to potential links between research involvement and community-engaged research outcomes, warranting longitudinal evaluation of the underlying causal pathways.

Increased research involvement was significantly associated with improved patient and community partnerships (i.e., partnership quality, research engagement, trust in medical researchers, and partnership recommendation). Importantly, these outcomes did not differ by race. The association with trust in medical researchers is particularly relevant given that confidence in scientists to act in the public’s best interests has declined among U.S. adults, from approximately 87% in 2020 to 76% in 2024.^32^ This is consistent with the present study’s findings, in which trust emerges from sustained partnership instead of episodic consultation,^33^ suggesting that greater involvement may help build trust in patients and communities where mistrust of research institutions has historically limited participation.^34^ These outcomes were associated with being a patient or community partner as well as the extent of these partnerships. Together, these findings indicate that patient and community involvement may strengthen a partnership’s quality and its long-term sustainability. One potential explanation is that increased involvement provides patient and community partners with greater opportunities to actively contribute to research activities, fostering a sense of respect, trust, and shared ownership.^12,14^

Associations between research involvement and partnership quality were primarily driven by roles in which participants had a more active impact. Participants involved in grant writing and shared governance may have had increased opportunities to influence a study’s priorities and decision-making rather than simply providing feedback. Notably, participation on a community advisory board was not associated with any of the examined outcomes; however, shared governance was associated with each outcome. While community advisory boards are useful, they often face challenges in recruitment, trust, and retention^35,36^ and may be viewed as tokenistic.^37^ They nonetheless remain among the most common structures for engaging communities in research,^38^ a prevalence that may reflect longstanding funder expectations^39^ and the relative ease of convening an advisory group within short timelines.This implies that the value of advisory structures may not depend solely on their existence, but on the quality of engagement they enable.^40^

Patient-centered research may benefit from enhancing shared governance in research as a means for patient and community engagement. This finding aligns with several frameworks stating that shared governance may enable more equitable participation and address unbalanced power dynamics between researchers and community partners.^41,42^ For example, Arnstein’s Ladder of Citizen Participation places community partnership over consultation.^43^ The present findings suggest that striving for the higher rungs of this ladder, such as through shared governance, may be associated with improved partnerships. Shared governance structures may therefore be associated with improved partnerships and offer a more sustainable approach.

Beyond governance-related activities, involvement in recruitment and data collection was also associated with improved partnership outcomes. These activities may provide patients and communities with direct opportunities to utilize their lived experiences and observe their contributions’ impact. Such visibility may reinforce partners’ sense of shared ownership of the research process, suggesting that meaningful engagement includes ensuring that partners’ roles involve substantive contributions to study implementation.

Together, these findings demonstrate the need for meaningful patient and community engagement throughout the research process. Many types of research involvement that were associated with individual outcomes were also associated with all four outcomes. While these outcomes each measure different aspects of patient and community partnerships, they may be related and reinforce one another.

This study’s cross-sectional nature enables limited causal inference. Participants more involved in research may perceive increased partnership quality, trust, and engagement; however, individuals who already have more positive perceptions of research may also be more likely to pursue greater research involvement. Longitudinal studies are needed to clarify whether increased research involvement precedes improved partnership outcomes. Additionally, this study consisted of patient and community partners. Therefore, this study may not be generalizable to individuals who have not participated in research partnerships. Finally, all measures were self-reported and may be subject to social desirability bias. Still, this study is among the first to examine how research involvement is associated with the quality of partnerships with patients and community members.

## Conclusion

In this national study of patient and community research partners, the extent of partners’ involvement in research was significantly associated with improved partnership outcomes.

Notably, not all forms of research involvement are associated with the same outcomes. Roles involving equal decision-making authority or those that create more visible impact for patient partners, such as shared governance, grant writing, recruitment, and data collection, were consistently associated with improved partnership outcomes. In contrast, such associations were not observed for consultative roles, such as community advisory boards. These findings suggest that future patient-centered outcomes research should involve patient and community partners throughout the research process and focus on active roles that foster shared ownership between researchers and community partners.

## Acknowledgments

We are grateful to the patient and community research partners for their time and contributions.

## Funding

This work was supported by the Patient-Centered Outcomes Research Institute (GC11046-01).

## Data availability

The data that support this study’s findings are available from the corresponding author upon reasonable request.

## Declarations

### Ethics approval and consent to participate

All participants provided informed consent, and all study procedures were approved by the Dartmouth Health Institutional Review Board.

### Consent for publication

Not applicable.

### Competing interests

None declared.

## Notes

### Competing Interest Statement

The authors have declared no competing interest.

### Author Declarations

All study procedures were approved by the Dartmouth Health Institutional Review Board.

### Summary of Updates

Author name updated to include a middle initial.

