## Appendix A for "Research Involvement and Partnership Outcomes Among Patient-Centered Outcomes Research Partners"

**Appendix A: Survey Items**

Quality of Patient-Centered Outcomes Research Partnerships (QPCOR):

(1)	I had a clear understanding of the purpose of the study.

(2)	I felt listened to.

(3)	I feel prepared to be an equal partner in the research study.

(4)	Researchers were knowledgeable about people like me or were willing to learn about people like me.

(5)	I believe that I had choices in how I could be a part of the research study.

(6)	I felt accepted by all members of the research study team.

(7)	Researchers used language that was consistent with my values and culture.

(8)	Both community members and researchers are thinking of ways we can continue to work together in the future.

(9)	I felt comfortable engaging with the members of the research study team.

(10)	I felt my views were incorporated into the research study.

Brief Research Engagement Tool:

(1)	The focus is on problems important to the community.

(2)	All partners assist in establishing roles and related responsibilities for the partnership.

(3)	Community-engaged activities are continued until the goals (as agreed upon by all partners) are achieved.

(4)	The partnership adds value to the work of all partners.

(5)	The team builds on strengths and resources within the community or patient population.

(6)	All partners’ ideas are treated with openness and respect.

(7)	All partners agree on the timeline for making shared decisions about the project.

(8)	The partnership’s processes support trust among all partners.

(9)	Mutual respect exists among all partners.

Trust in Medical Researchers Scale:

(1)	To get people to take part in a study, medical researchers usually do not explain all of the dangers about participation.

(2)	Participants should be concerned about being deceived or misled by medical researchers.

(3)	Usually, researchers who make mistakes try to cover them up.

(4)	Medical researchers act differently toward minority subjects than toward White subjects.

(5)	Medical researchers unfairly select minorities for their most dangerous research studies.

(6)	Some medical research projects are secretly designed to expose minority groups to diseases such as AIDS.

(7)	Medical researchers are generally honest in telling participants about different treatment options available for their conditions.

(8)	Usually, medical researchers tell participants everything about possible dangers.

(9)	All in all, medical researchers would not conduct experiments on people without their knowledge.

(10)	Most medical researchers would not lie to people to try to convince them to participate in a research study.

(11)	In general, medical researchers care more about doing their research than about the participants’ medical needs.

(12)	Researchers are more interested in helping their careers than in learning about health and disease.
